# Comprehensive, Transparent, and Fair Machine Learning Models for Hypertension Risk Prediction: Benchmarking With Framingham, External Validation, Individual-Level Analysis, and Equitable Clinical Utility

**DOI:** 10.1101/2025.09.03.25334979

**Authors:** Parsa Amirian, Mahsa Zarpoosh

## Abstract

**Background:** Hypertension (HTN) is a leading, yet often underdiagnosed, cause of cardiovascular diseases worldwide. While clinical risk scores like the Framingham Risk Score (FRS) are commonly used, their limited capacity to capture complex risk patterns and their poor generalizability hinder optimal prediction and prevention in resource-limited settings.

**Methods:** We developed and rigorously validated a comprehensive suite of machine learning (ML) models, including tree-based, linear, kernel, neural network, and ensemble classifiers for incident HTN prediction using data from our internal cohort (n=8,054, Iran). All models were benchmarked against the FRS. External validation was conducted on the NHANES cohort (n=6,266, USA), employing harmonized features. We systematically assessed model performance across multiple metrics (ROC AUC, PR AUC, F1, and Brier), evaluated calibration, clinical decision benefit, and learning curves. We conducted extensive subgroup and fairness analyses by sex, education, and socioeconomic status. Interpretability was ensured via SHAP values and permutation importance. Additionally, individualized counterfactual analyses, bootstrap prediction intervals, and patient-level risk vignettes were provided.

**Results:** ML models, especially our ensembles and LightGBM, significantly outperformed FRS in discrimination (mean ROC AUC improvement up to 0.04, 95% CI not crossing zero), with robust generalizability confirmed in external validation. All models demonstrated minimal performance disparities across demographic and socioeconomic subgroups, and SHAP analyses identified SBP, age, and BMI as consistently strong predictors. Individualized risk vignettes illustrated the clinical nuance of ML predictions compared to categorical clinical scores.

**Conclusion:** This open, reproducible study demonstrates that modern, interpretable ML models provide significant and equitable improvements over standard clinical risk scores for HTN prediction, with strong external validity and clinical utility. An online risk calculator is provided to facilitate real-world deployment.

## Introduction

Hypertension (HTN) is one of the most significant public health challenges worldwide. Approximately 1.28 billion people are living with HTN, and less than half are aware of their condition (1). HTN is a leading risk factor for cardiovascular diseases (CVDs), causing stroke, myocardial infarction, heart failure, and kidney failure. Annually, deaths linked to HTN reach 106.3 per 100,000 persons. Although HTN is easily treatable and end-organ damage can be prevented by antihypertensive drugs, lack of proper diagnosis and the fact that two-thirds of the cases are in the middle and low-income countries, developing accessible diagnostic tools is a critical public health priority (2,3).

There are several questionnaire-based HTN risk scores; the Framingham Risk Score (FRS) is among the most validated ones (4). FRS utilizes several fundamental risk factors, including age, sex, systolic blood pressure (SBP), and diastolic blood pressure (DBP), and then translates these continuous variables into point scores. The FRS is easy to calculate and interpret without specialized tools and has been extensively validated across multiple populations, with its findings integrated into guidelines. Nonetheless, the FRS was developed from a predominantly white cohort in the mid-20th-century US and cannot capture complex patterns in data, nonlinear relationships, or interactions. In this study, we used the FRS as a standard clinical benchmark against which to compare modern Machine Learning (ML) risk calculators.

ML models, including random forest (RF), support vector machines (SVM), and extreme gradient boosting (XGBoost), have shown better accuracy over traditional risk scores; nevertheless, prior studies often miss external validation, fairness assessment, and rarely explain how their ML models drew the results. Furthermore, calibration and clinical utility are usually absent in prior studies (5,6). For ML models to consistently demonstrate outperformance and establish their superiority over traditional risk scores, such as FRS, they must be transparent, truly externally validated, explainable, and fair across different population groups (7).

In this study, we aimed to perform head-to-head comparison between all prominent ML families including tree-based models (LightGBM, XGBoost, Gradient Boosting (GB), Decision Tree (DT), and RF), linear models (logistic regression (LR)), kernel-based models (SVM), neural network models (neural net (NN)), and three ensembles (stacking, voting, and weighted) and one hybrid ML-FRS (meta-ensemble) against well-established FRS clinical risk score and externally validate the results in an ethnically different cohort. Furthermore, we assessed the fairness of our models across diverse subgroups, including individuals with different socioeconomic status (SES), educational backgrounds, and both males and females. To guarantee the models are understandable and transparent, we calculated Shapley additive explanations (SHAP) values for each model. Moreover, we utilized case vignettes to assess ML models at the individual level.

We provide an open, end-to-end pipeline, complete with an online risk calculator, for clinical deployment. Such comprehensive analytic transparency and clinical utility in developing robust, transparent, and globally applicable HTN risk prediction ML models represent a significant advancement over traditional approaches or previous ML risk prediction papers.

## Methods

### Study Design

Initially, we utilized the data of 9580 participants of the Ravansar Non-Communicable Disease (RaNCD) prospective cohort as our internal dataset. RaNCD is a part of Prospective Epidemiological Research Studies in Iran (PERSIAN) mega cohort (8). Entrants’ age is between 35 and 70, and they reside in both urban and rural areas of Ravansar county in the western part of Kermanshah province. The data is acquired through various means, including anthropometry, physical examination, questionnaires, and laboratory tests. The dataset was mostly complete, with a few imputations needed. Only 11 observations were missing, all in the categorical variables. We used the mode imputation method for these missing values, and all 11 instances were assigned the category “negative.” No systematic patterns were identified in the missing data, indicating that the missingness was most likely completely at random. To address outliers among the continuous variables, we used a combined normalization and clipping approach before model fitting. All continuous variables were first standardized (z-transformed), and then values exceeding three standard deviations from the mean (z-score > 3 or <-3) were clipped to ±3. This procedure was consistently applied within each fold of the cross-validation and test splits; therefore, scaling and clipping statistics were derived only from the respective training data, avoiding data leakage. The prevalence of HTN in the internal cohort was 1526 cases; we removed those participants from the study as the first step and started the analysis with 8054 entrants (see Figure 1 for study overview). For external validation of our HTN prediction models, we employed the publicly available National Health and Nutrition Examination Survey (NHANES) dataset, which is accessible through Kaggle (https://www.kaggle.com/datasets/cdc/national-health-and-nutrition-examination-survey). HTN was defined as a SBP of 140 mmHg or higher and/or a DBP of 90 mmHg or higher, determined from two or more measurements obtained by the study personnel or attending physicians. None of the included participants had SBP or DBP readings indicative of HTN on their initial measurement. The NHANES dataset comprised 9813 observations. To ensure that there are enough observations for rigorous external validation, we did not exclude any NHANES entrant due to missing data. Across mapped continuous variables, 2316 instances were missing, so we used the mean imputation method to fill these gaps in the dataset. The “Educational years” categorical variable had 680 missing instances; we utilized the proportional imputation method for these missing values. The status of HTN variables in the NHANES dataset had 3547 missing values. We omitted those with missing HTN status in the NHANES and performed the external validation on 6266 subjects. To maintain consistency and prevent data leakage, continuous variables in NHANES were standardized and clipped based on the parameters (mean, standard deviation (SD)) derived solely from the internal training cohort.

**Figure 1.**
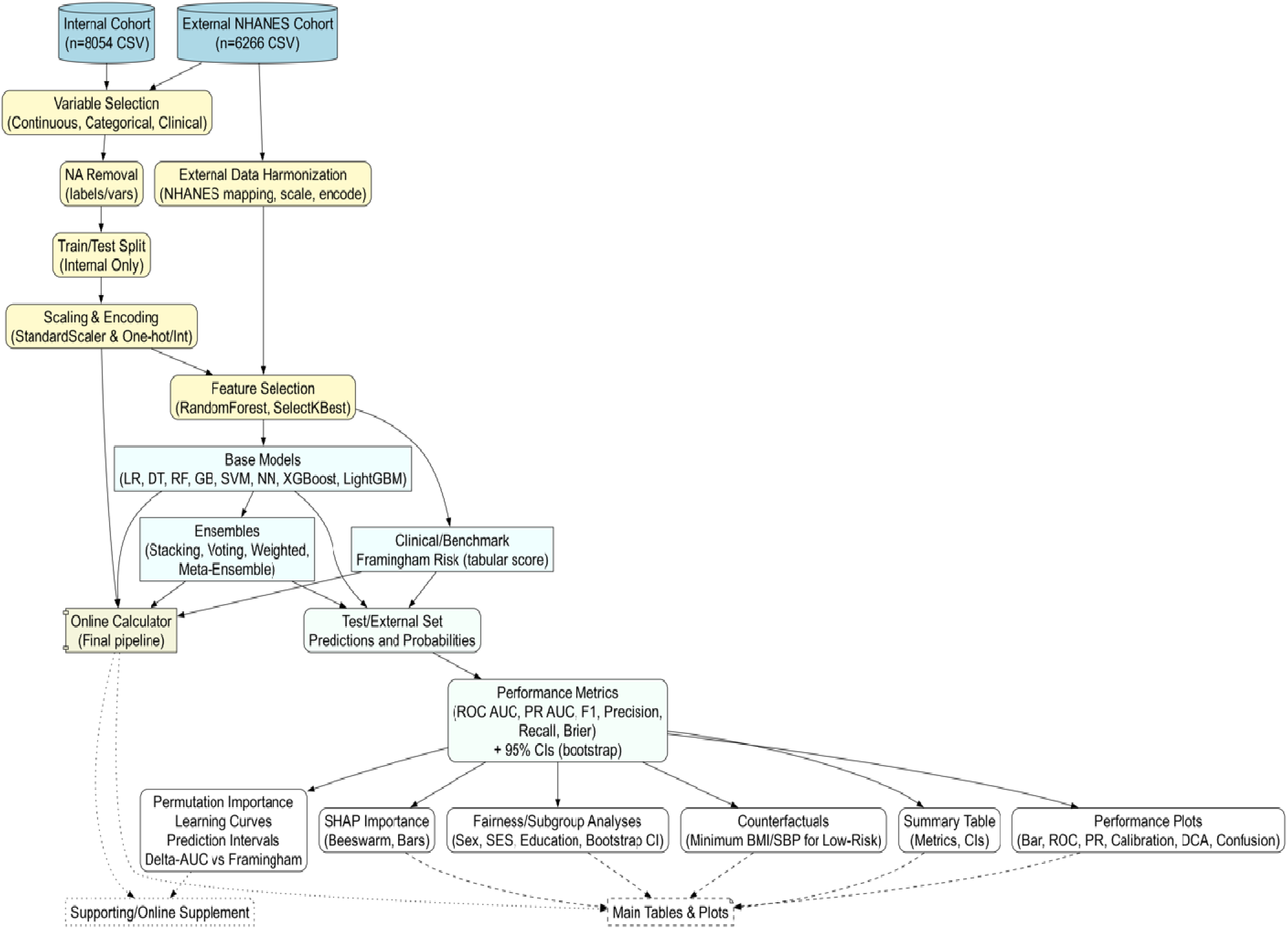
Study Design and Pipeline Overview.

### Features

We began by assembling a comprehensive set of candidate predictors, the internal dataset included 18 continuous variables (age, metabolic equivalent (MET), fasting blood sugar (FBS), wrist circumference, waist to hip ratio (WHR), protein intake, carbohydrate intake, total energy intake, educational years, sleep duration, SBP, DBP, triglyceride (TG), cholesterol, high-density lipoprotein (HDL), low-density lipoprotein (LDL), body mass index (BMI), and socioeconomic status quintile (SESq)). Continuous variables underwent standardization (z-score normalization) based on the training set, with test and external validation sets transformed using training-derived parameters. After standardization, values exceeding ±3 standard deviations were clipped to (−3, 3) to mitigate the potential influence of outliers. One-hot encoding was applied to multi-category non-hierarchical variables. Twenty categorical variables were also included; all binary coded 0 for absence/no, 1 for presence/yes. The variables are as follows: residence (urban or rural), marital status (single or not single), ), history of gastroesophageal reflux disease (GERD), history of blood in stool, history of statin prescription, diagnosed with osteoporosis (yes or no), history of rheumatoid arthritis (RA), history of weight loss, history of muscle weakness, history of anti-triglyceride drug prescriptions, alcohol consumption (yes or no), history of type 2 diabetes mellitus (T2DM), history of cardiovascular diseases (CVD), familial history of T2DM, familial history of HTN, familial history of cardiac diseases, familial history of myocardial infarction (MI), familial history of stroke, T2DM incidence, and smoking status (current smoker or not current smoker). In each classifier’s pipeline, model-based or univariate feature selection was applied based on tuned parameters to improve parsimony.

We were able to confidently map seven variables across NHANES and our internal cohort (Table 1 of S1 file). The variables were carefully inspected to choose the most similar features in the two cohorts. Seven variables, including age, educational years, SBP, DBP, BMI, total energy intake, and total protein intake, were systematically mapped.

**Table 1.** Demographic, clinical, and lifestyle characteristics comparison between HTN and non-HTN subjects in (a) internal cohort and (b) NHANES 2013-2014 cohort.

| Characteristics | Internal cohort |  |
| --- | --- | --- |
| | Mean ± SD or N (percentage) | |
|  | Non-HTN (N = 7193) | HTN (N = 861) |
| Age (years) | 45.99 ± 7.90 | 49.73 ± 7.76 |
| Sex |  |  |
| Male | 3638 (50.58) | 324 (37.63) |
| Residence type |  |  |
| Urban | 4328 (60.17) | 539 (62.60) |
| Marital status |  |  |
| Single | 531 (7.38) | 76 (8.83) |
| MET | 41.37 ± 8.43 | 40.09 ± 7.36 |
| FBS | 94.99 ± 26.86 | 100.54 ± 35.57 |
| Statin use |  |  |
| Yes | 142 (1.97) | 37 (4.30) |
| Anti-TG drug use |  |  |
| Yes | 55 (0.76) | 11 (1.28) |
| Wrist circumference | 17.09 ± 1.38 | 17.13 ± 1.41 |
| SES quantiles |  |  |
| 1 | 1348 (18.74) | 179 (20.79) |
| 2 | 1366 (18.99) | 198 (23.00) |
| 3 | 1461 (20.31) | 177 (20.56) |
| 4 | 1477 (20.53) | 164 (19.05) |
| 5 | 1541 (21.42) | 143 (16.61) |
| Alcohol |  |  |
| Yes | 390 (5.42) | 24 (2.79) |
| WHR | 0.93 ± 0.06 | 0.95 ± 0.05 |
| T2DM |  |  |
| Yes | 471 (6.55) | 99 (11.50) |
| CVD |  |  |
| Yes | 470 (6.53) | 61 (7.08) |
| T2DM (FH) |  |  |
| Yes | 1814 (25.22)) | 221 (25.67) |
| HTN (FH) |  |  |
| Yes | 3365 (46.78) | 436 (50.64) |
| Daily protein (gr) | 93.71 ± 37.57 | 86.36 ± 34.19 |
| Daily carbohydrate (gr) | 421.87 ± 151.33 | 398.15 ± 147.10 |
| Daily energy (kcal) | 2760.99 ± 971.66 | 2582.05 ± 918.86 |
| Education years | 5.86 ± 4.82 | 4.43 ± 4.61 |
| Sleep duration (hours) | 7.10 ± 1.22 | 6.98 ± 1.20 |
| SBP (mmhg) | 103.56 ± 12.35 | 111.65 ± 12.05 |
| DBP (mmhg) | 67.45 ± 7.80 | 71.34 ± 8.30 |
| TG | 134.33 ± 79.85 | 147.11 ± 87.83 |
| Total cholesterol | 184.23 ± 37.34 | 189.48 ± 40.18 |
| HDL | 46.42 ± 11.36 | 45.85 ± 11.37 |
| LDL | 110.94 ± 30.85 | 114.21 ± 33.57 |
| BMI | 27.18 ± 4.59 | 28.76 ± 4.46 |
| Smoking |  |  |
| Current | 909 (12.64) | 91 (10.57) |

| NHANES cohort (2013-2014) |  |  |
| --- | --- | --- |
| Characteristics | Mean ± SD or N (percentage) |  |
|  | Non-HTN (N = 4148) | HTN (N = 2118) |
| Age (years) | 39.24 ± 17.52 | 58.40 ± 15.87 |
| SBP | 117.18 ± 14.04 | 130.64 ± 18.93 |
| DBP | 68.02 ± 11.53 | 70.62 ± 13.95 |
| Daily energy (kcal) | 2175.35 ± 1006.68 | 2009.76 ± 927.56 |
| BMI | 27.54 ± 6.66 | 31.06 ± 7.54 |
| Daily protein (gr) | 83.86 ± 47.63 | 76.32 ± 40.51 |
| Education quantile |  |  |
| 1 | 301 (7.26) | 185 (8.73) |
| 2 | 538 (12.97) | 311 (14.68) |
| 3 | 889 (21.43) | 523 (24.69) |
| 4 | 1272 (30.67) | 680 (32.11) |
| 5 | 1148 (27.68) | 419 (19.78) |

### Framingham Risk Score

All ML models need a clinical benchmark for comparison. To provide a rigorous clinical reference for model performance, we implemented the FRS as a benchmark. In our study, we computed the FRS for each participant in the internal cohort using the published point-based algorithm (Table 2 of S1 file) (4). The included variables were age, SBP, BMI, familial history of HTN, and smoking status. To enable robust comparison with the ML models, we constructed a logistic regression model using the FRS as the sole predictor for incident hypertension within our test set. The construction allowed for recalibration of the risk score to the characteristics and outcome distribution of our study population. The logistic regression-derived predicted probabilities were used as continuous predictions for all statistical evaluations (see the pipeline, part 8). The final Framingham score for each participant was calculated by summing the assigned points across all risk factors; moreover, for comparison of binary classification metrics, a pre-specified score threshold of ≥ 8 points (as per standard practice) was used to represent risk predictions.

**Table 2.**
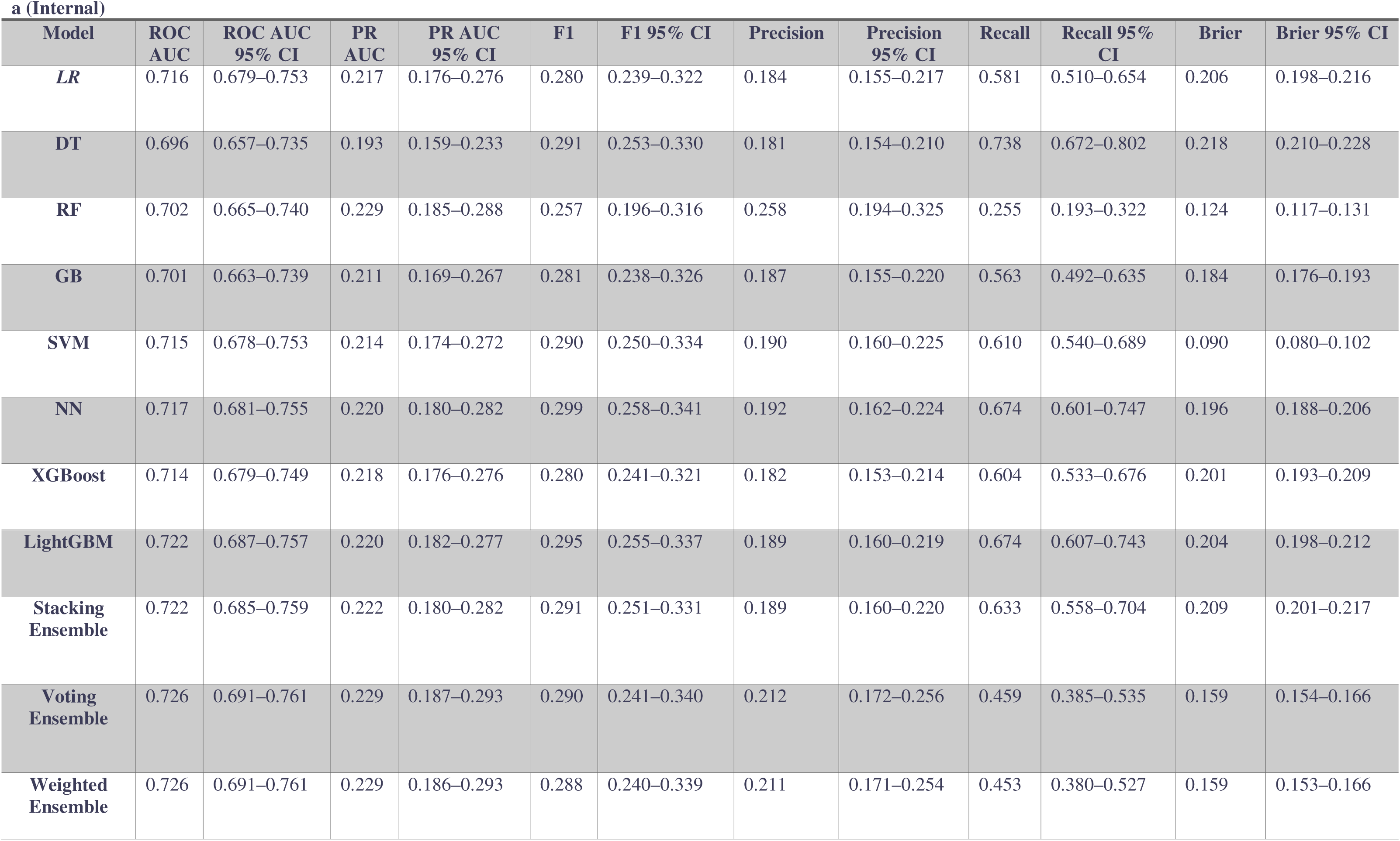
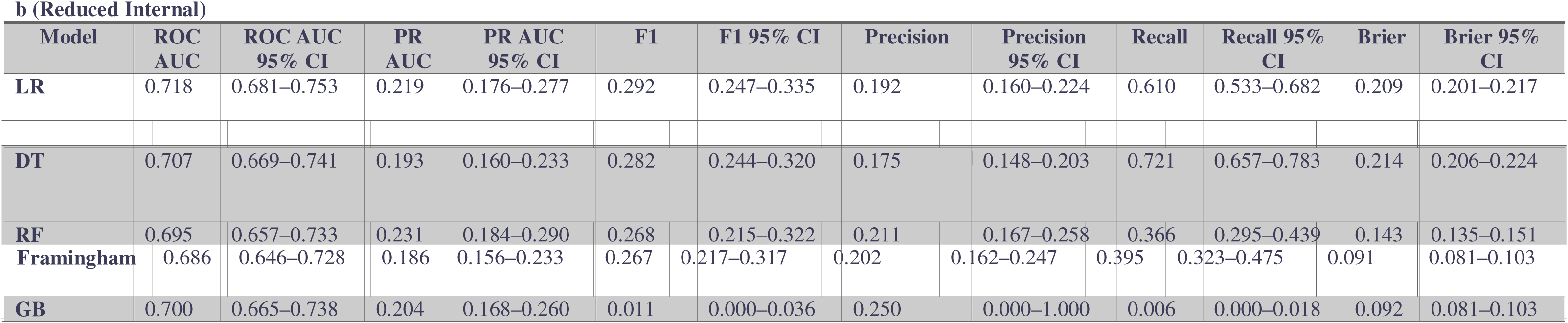
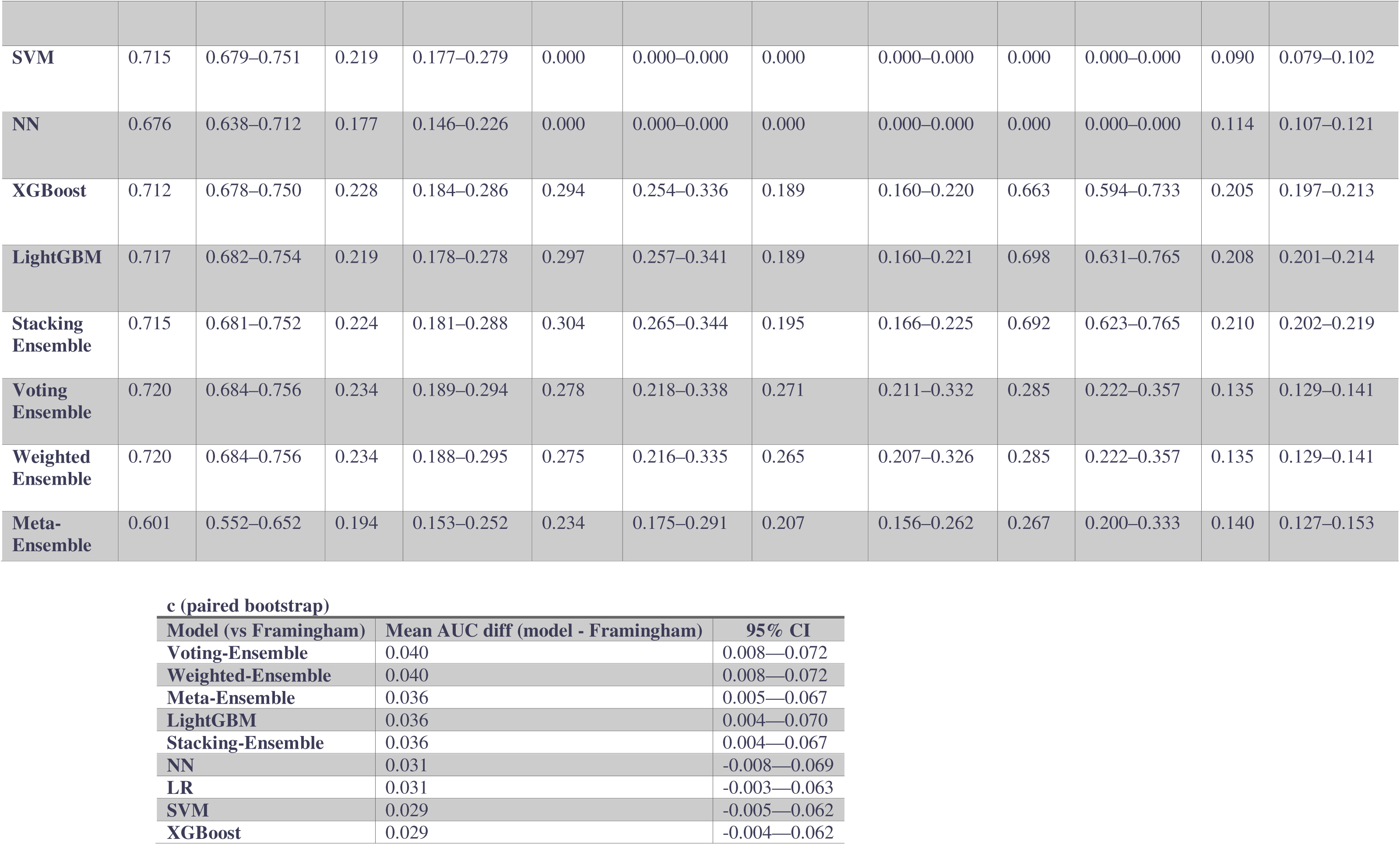
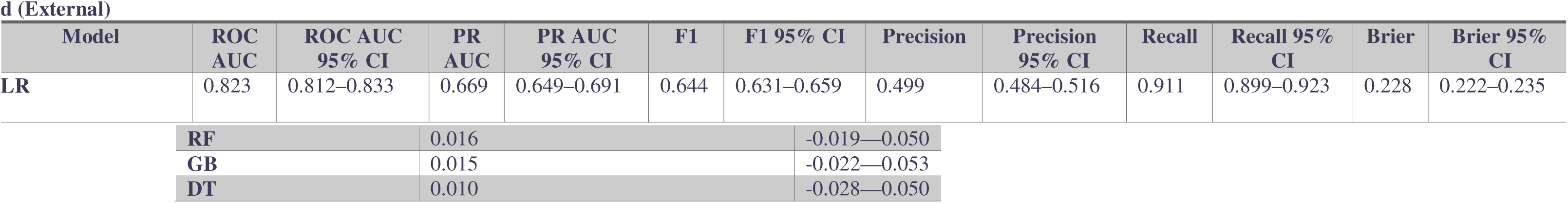
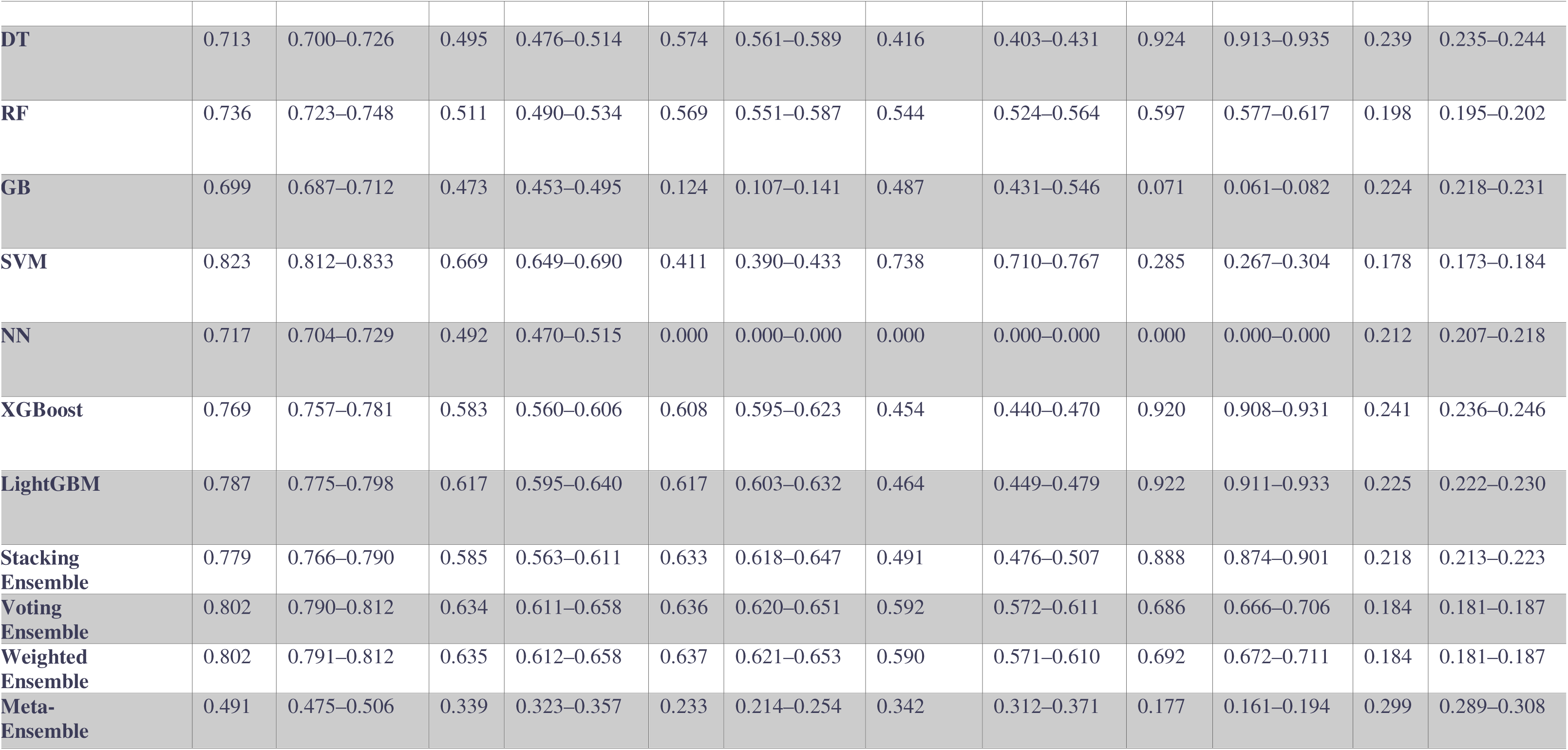
Performance metrics of ML models across internal, reduced internal, and external test sets. (a) Internal test set results showing ROC AUC, Precision-Recall AUC, F1 score, Precision, Recall, and Brier score with 95% confidence intervals. (b) Performance on the reduced internal feature set. (c) Paired bootstrap comparison of mean ROC AUC differences between each model and Framingham score with corresponding 95% confidence intervals. (d) External validation results reporting ROC AUC, Precision-Recall AUC, F1 score, Precision, Recall, and Brier score with 95% confidence intervals.

### Data Partitioning and Handling Class Imbalance

All data pre-processing, model development, evaluation, and reporting were performed in accordance with the Transparent Reporting of a multivariable prediction model for Individual Prognosis or Diagnosis (TRIPOD) guidelines (10). The internal cohort was randomly divided into training and internal holdout (testing) sets using stratified random sampling to preserve the proportion of HTN incidence across both partitions. Specifically, 80% of samples were allocated to the training set and 20% to the internal test set. All models were fitted exclusively on the training subset. The exclusiveness included the calculation and fitting of transformers, resampling strategies, and feature selection methods. To allow for reproducibility, we fixed the random state to 42.

HTN incidence was relatively imbalanced in our internal cohort. To reduce the risk of bias toward the majority class, we systematically addressed class imbalance using the following model-specific strategies. Class weighting was used for the majority of the models, except for NN, for which we used SMOTE (Synthetic Minority Over-Sampling Technique) in the training set. All ensembles inherited class imbalance strategies from their components; furthermore, FRS was a reference and was not fitted or adjusted for imbalance. Additionally, no resampling or weighting was ever applied to test or external validation sets.

### Machine Learning Models

We had a broad-spectrum view about choosing ML models for predicting HTN incidence. We included the simplest (e.g., LR) to the most sophisticated (ensembles) ML classifiers in this study. The following classifiers were included: LR, LightGBM, XGBoost, GB, DT, RF, SVM, NN, soft voting ensemble, weighted soft voting ensemble, stacking ensemble, and meta-ensemble. To optimize model performance, hyperparameters for each model were selected using random search within pre-specified ranges based on initial experimentation. Five-fold stratified cross-validation on the training set was used to estimate the performance of each parameter configuration. The complete hyperparameter search space for each classifier is detailed in Table 3 of the S1 file. Four ensemble approaches were utilized to combine information from first-line classifiers. Our first implemented ensemble was a stacking ensemble using a logistic regression meta-learner, combining the out-of-fold predicted probabilities of all individual machine learning models. Next, a soft voting ensemble was created by averaging the predicted probabilities of all base classifiers for each sample; furthermore, a weighted soft voting ensemble was also constructed by assigning each base model a weight proportional to its mean cross-validated Area under the receiver operating characteristic curve (ROC AUC) on the training set. Each model’s probability was multiplied by its respective weight, and then the sum was used as the final prediction for each sample. Lastly, we created a meta-ensemble by training a logistic regression model on a feature set that included the FRS and the predicted probabilities from base models.

The meta-ensemble integrated both clinical and machine learning predictors, allowing it to optimize their relative contributions to risk estimation. For a head-to-head comparison of models between ML models in the internal and external cohorts, reduced ML models are used, which rely only on the mapped features available in both cohorts.

### Performance evaluation

Following best practices in clinical ML, we conducted an extensive performance evaluation of all models using internal and external validation cohorts. For each model, we assessed several metrics on the internal hold-out test set and external dataset, namely ROC AUC, Area under the precision recall curve (PR AUC), and classification metrics including F1 score, precision, and recall using the hard threshold of 0.5. Moreover, we reported the agreement between predicted probabilities and observed outcomes using calibration curves and the Brier score. To assess clinical utility across decision thresholds, we computed net benefit curves for all models, plotting net benefit across a range of threshold probabilities (0.01–0.99).

Interpretability and feature relevance were explored by permutation importance analysis; for each model, we report the decrease in test-set ROC AUC when each top feature is randomly permuted, to identify which features are most critical to model performance. Additionally, for each model, global Shapley Additive exPlanations (SHAP) importance values were computed, and barplots and beeswarm plots were drawn. To assess model learning behavior and possible overfitting/underfitting, learning curves were generated per model, which plot training and cross-validation ROC AUC as a function of training sample size. To assess the fairness of the models between males and females, we plotted the fairness gap of each model for all metrics for their respective 95% CIs. Subgroup analysis was also conducted to evaluate the performance across subpopulations. We stratified the test set by sex, SESq, and education level. For each subgroup and model, we report ROC AUC, PR AUC, F1, Brier, precision, and recall, along with their respective 95% CIs. Calibration curves and decision curve analysis (DCA) were also visualized for the sex category. All metrics and interval estimates were benchmarked against the FRS as a clinical baseline. In addition, the statistical significance of discrimination differences between models and Framingham was assessed using paired bootstrap estimates of ROC AUC differences.

### Case vignettes

To maximize the clinical message, we utilized the case vignette approach. We selected 10 representative test-set patients. For each, we calculated the predicted risk of hypertension across all models using the patient’s actual features, examined risk differences, and used counterfactual analyses to estimate the minimum decrease in BMI and SBP required to reduce estimated risk below a 0.2 threshold. We visualized the predicted absolute risk of these entrants across the best models by spider plots; furthermore, we showed risk trajectories for BMI and SBP through line plots, showcasing how risk responds as these modifiable factors improve. Bootstrap prediction intervals (PI) were plotted for 10 case vignettes to assess the model’s uncertainty further. For each model, we refitted the pipeline on bootstrap samples and estimated the 95% empirical interval (2.5–97.5th percentile) for each prediction.

### Reproducibility and Software

All analyses were performed in Python 3.12.7, using scikit-learn 1.6.1, SHAP 0.47.2, pandas 2.2.3, NumPy 1.26.4, Seaborn, Matplotlib, Graphviz for visualization, and STATA 17 (StataCorp. 2025. Stata Statistical Software: Release 17. College Station, TX: StataCorp LLC.) for the initial statistical observation. To follow the open science policy and complete transparency, the entire code, the external cohort’s data, and the internal mock dataset will be made public immediately upon acceptance or publication in a GitHub repository. To further incorporate this study into clinical practice, we developed a web-based clinical decision-support application based on the externally validated models. The calculator is publicly available at (https://htn-calculator.onrender.com/). The app is open-sourced, and the code is available at (https://github.com/ParsaMD/HTN_calculator).

## Results

### Cohort characteristic

The current study comprises 8054 internal entrants with a mean age ± standard deviation (SD) of 46.39 ± 7.97. The cohort included 4092 (50.81%) females and 3962 (49.19%) males; furthermore, 861 (10.69%) participants developed HTN during the follow-up period (Table 1a). The mean SBP in the internal cohort was 104.43 ± 12.57, and the mean DBP was 67.87 ± 7.95. Our external validation cohort (NHANES 2013-2014) comprised 6266 participants with the mean age ± SD of 45.71y ± 19.25 (Table 1b), the NHANES cohort featured 2218 (33.80%) HTN cases (prevalence cases). The mean SBP in the NHANES cohort was 121.73 ± 17.09, and the mean DBP was 68.90 ± 12.46. The full characteristics of the included 14420 participants in the analysis are shown in Table 1.

### Internal models’ performance

Table 2 shows the results of all main (Table 2a) and reduced (Table 2b) ML models on the internal test set. As shown in Figure 2a, all models demonstrate moderate to strong discrimination ability on the internal test set using the complete feature set. Ensembles were the best performers. Our best performance was achieved by our voting and weighted ensembles, with ROC AUC (95% CI) of 0.72 (0.69–0.76 and our meta-ensemble also performed well (0.72, 0.68–0.76). In the base models, LightGBM performed exceptionally well and was comparable with ensembles, with an ROC AUC of 0.72 (0.68–0.75). The highest PR AUC was achieved by voting ensemble with PR AUC of 0.22 (0.18–0.29), the best F1 score was scored by NN with F1 of 0.29 (0.25-0.34). Furthermore, RF showed the highest quality of positive predictions (precision) at 0.25 (0.19-0.32), and DT had the highest recall rate at 0.73 (0.67-0.80). Finally, the lowest Brier score belonged to SVM at 0.09 (0.08-0.10); it is worth mentioning that Framingham also had a low Brier score at 0.09 (0.08-0.10). Reduced internal models, based on mapped parameters between internal and external cohorts, showed more or less the same results; for example, voting and weighted ensembles achieved the highest ROC AUC at 0.72 (0.68-0.75), the PR AUC at 0.23 (0.18-0.29). Notably, comparing reduced models, the stacking ensemble achieved the highest F1 score, and SVM achieved the lowest Brier score. In the reduced models, SVM and NN failed to predict a true positive case at a 0.5 threshold, thus they had zero F1 scores (Table 2b). Figure 2b illustrates model performance (ROC AUC, PR AUC, and F1) when limited to a reduced set of features. Confusion matrices, PR curves, and ROC curves of all internal main models are presented in Figure 1 of the S2 file and for the reduced models in Figure 4 of the S2 file.

**Figure 2.**
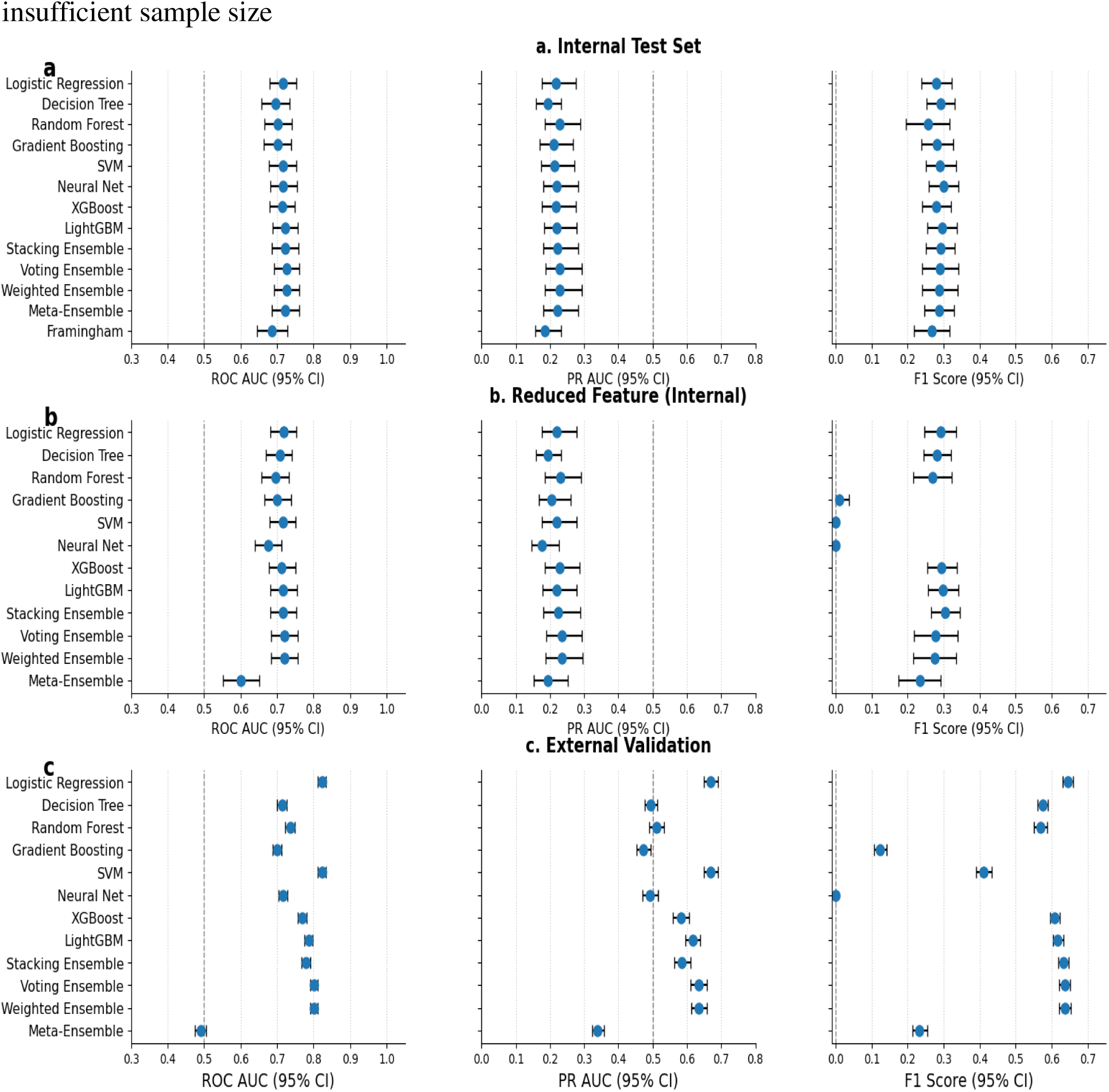
Performance comparison of ML models across three evaluation settings: (a) Internal test set with full feature set, (b) Internal test set with reduced features, and (c) External validation cohort. Each panel presents model performance metrics, including ROC AUC, PR AUC, and F1 score, with 95% confidence intervals.

**Figure 3.**
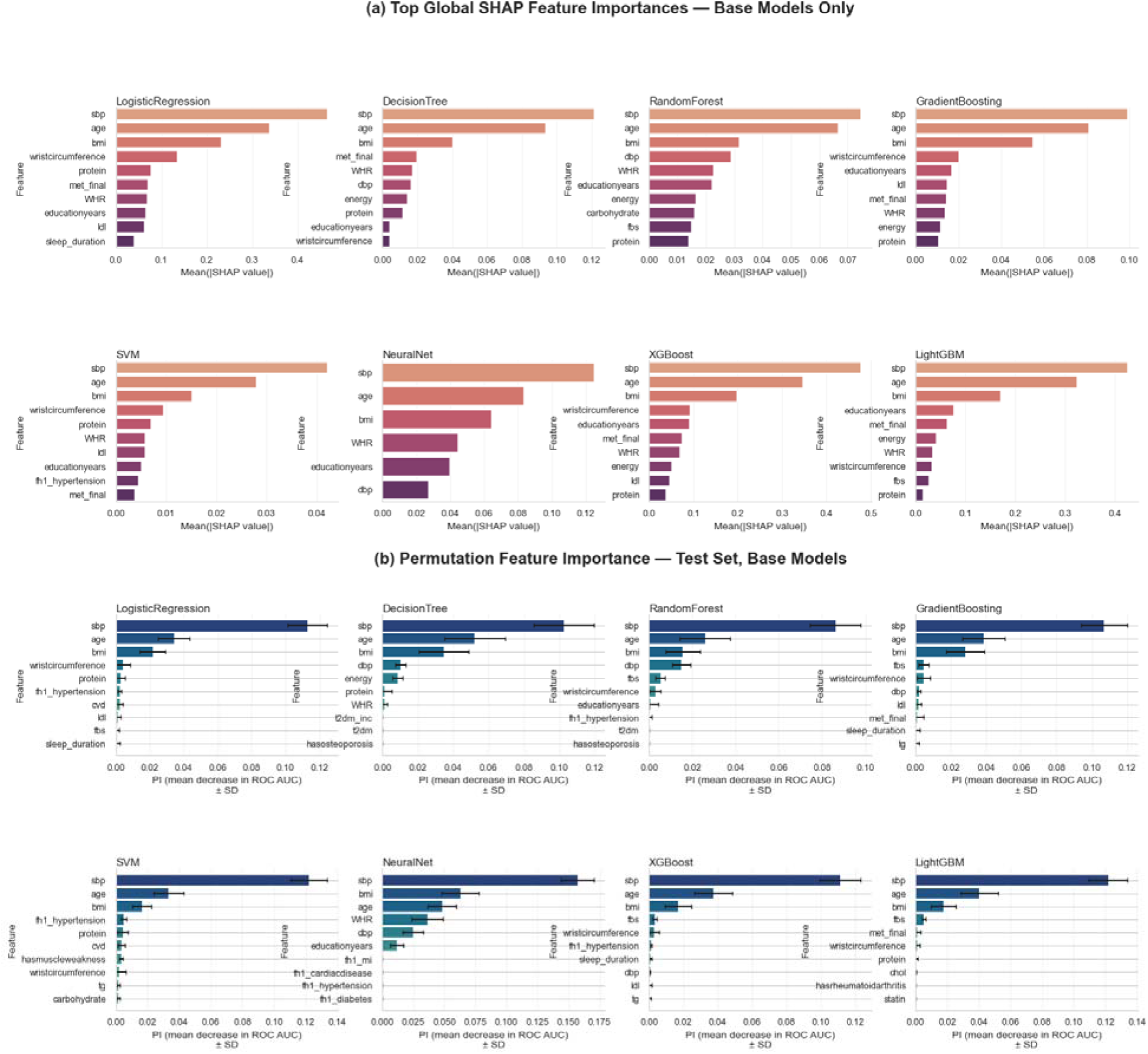
Comparison of feature importance rankings across base predictive models. (a) Mean absolute SHAP values representing global feature contributions, displayed for the top 10 features per model. (b) Permutation importance scores (mean decrease in ROC AUC ± standard deviation) computed on the test set for the top 10 features per model.

**Figure 4.**
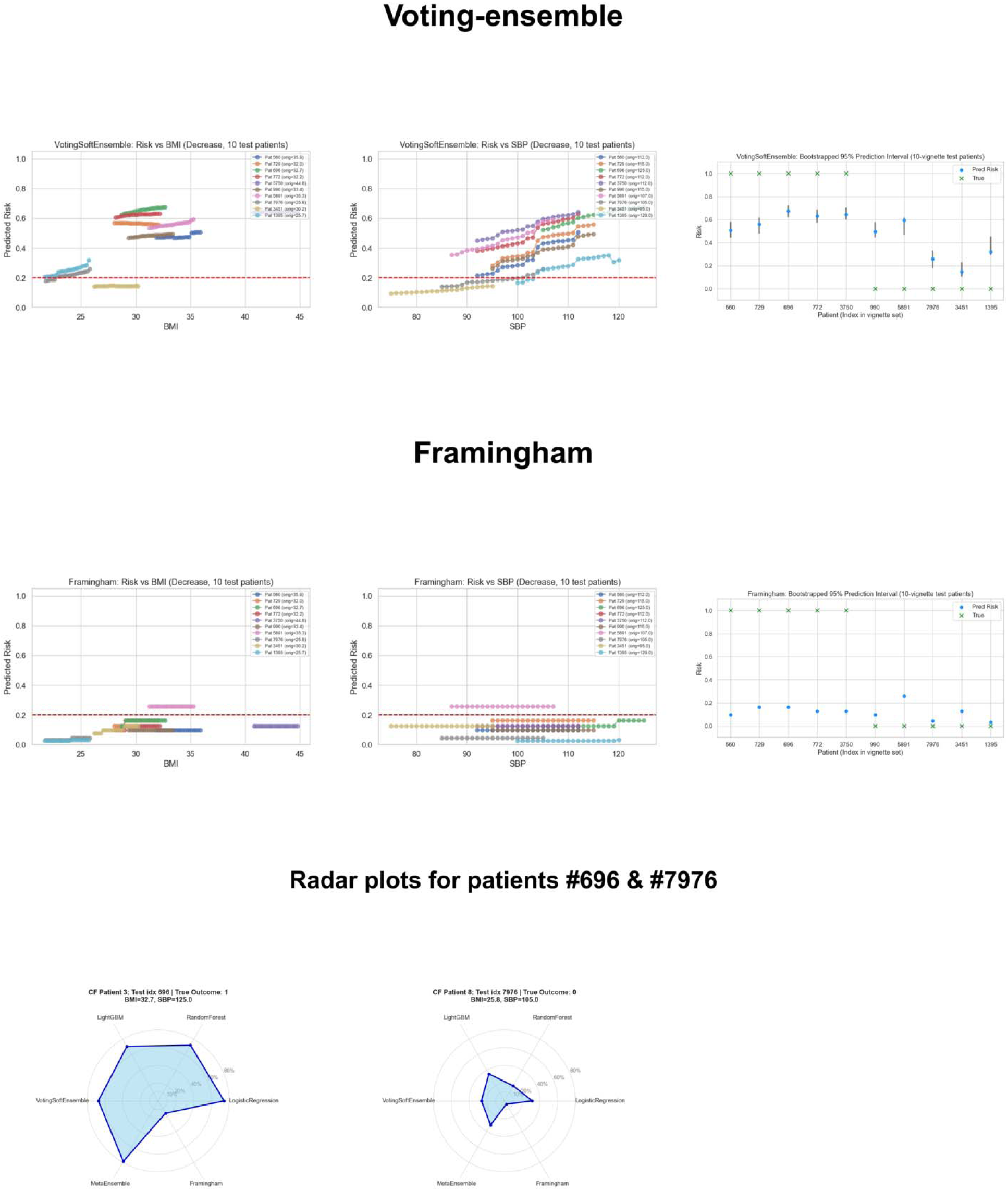
Individual-level counterfactual risk trajectories and 95% bootstrap prediction intervals for voting ensemble and Framingham models. Radar plots compare the predicted risks across six models for patients #696 & #7976.

### Internal models v. Framingham

For a head-to-head comparison between clinical benchmarks (Framingham) and ML models, a paired bootstrap test for mean differences in ROC AUC (with 95% confidence intervals) was performed. Five models, including all four ensembles and LightGBM, substantially outperformed the FRS, given the fact that their CIs did not cross zero. The voting ensemble and weighted ensemble models both showed the most significant increase in ROC AUC compared to the FRS, with a mean difference of 0.040 (0.008 to 0.072). Table 2c shows the full results of the head-to-head comparison.

### External validation

Generally speaking, ML models performed better ROC AUC-wise in the external cohort. For example, LR and SVM achieved ROC AUC of 0.82 (0.81-0.83), substantially higher (DROC 0.11) compared to their performance in the held-out internal test set. LR and SVM also had the best PR AUC at 0.66 (0.64-0.69). In the external cohort, the highest F1 score also belonged to LR at 0.64 (0.63-0.65); furthermore, the best precision belonged to SVM at 0.73 (0.71-0.76). DT and LightGBM scored an outstanding recall at 0.92 (0.91-0.93). All models had an acceptable Brier ranging from 0.17 to 0.24 in the external test set. Table 2d shows the full performance of ML models on the external cohort (NHANES). Performance on the external validation cohort, depicted in Figure 2c. Moreover, confusion matrices, PR curves, and ROC curves of all external models are presented in Figure 5 of the S2 file.

### Calibration & DCA

Calibration curves and DCA plots were drawn for all primary models, reduced internal, and external validation models. Figure 2 of the S2 file shows the calibration plots of all internal main models; by inspecting the plot, it is evident that almost all models fell below the diagonal, indicating that models generally overestimated the risk of HTN incident. Reduced internal models also had the same outcome, except for SVM, in 0.3 predicted probability; they were all below the diagonal, indicating the overestimation of the risk (Figure 6 S2 file). The result was not the same for external validation models; some models were above the diagonal. For example, GB, SVM, and NN were mainly above the diagonal, indicating underestimation of the true risk of HTN (Figure 7 S2 file). To be fully transparent and to have a clear clinical picture, we also calculated the net benefit, but due to low rates of HTN incidence and the extensive number of false positives, the net benefit was almost flat across all thresholds for all models (Figures 3,8, and 9 S2 file).

### Subgroup & Fairness analysis

All models, including Framingham, were evaluated for subgroup equity by sex, SES quintile, and education, with no evidence of major systematic performance disparities. However, performance variabilities were present, particularly in the smallest or most marginal groups. For example, discrimination for HTN incidence prediction was generally slightly higher in men compared with women, but absolute differences in ROC AUC were modest and CIs largely overlapping. ROC AUCs in women (n=801, prevalence 11.6%) ranged from 0.65 (DT, 0.59– 0.71) to 0.71 (stacking/meta ensemble & LR, 0.66–0.76). The full report of all metrics and their respective 95% CIs by each sex can be seen in Table 5 of the S1 file. All models’ ROC AUC (Figure 16), PR AUC (Figure 19), Brier (Figure 22), F1 (Figure 25), precision (Figure 28), and recall (Figure 31) plots by sex can be found in the S2 file.

Furthermore, to examine the fairness of our models between males and females, fairness gap analysis between men and women was performed. Generally, the gap was below 0.05 in almost all models for ROC AUC, PR AUC, F1, precision, and recall. 95% CIs mostly passed the zero threshold, indicating no systematic unfairness; nonetheless, the Brier score was unfair primarily to males, and only FRS and SVM’s CIs passed 0 in the Brier gap plot; additionally, the recall gap in ensemble models was significant, they were unfair to males, and their CIs did not cross the zero threshold. The complete fairness analysis can be found in Figures 10-15 of the S2 file and Table 4 of the S1 file.

Across socioeconomic strata (SESq 1–5), model performance was largely consistent, with ROC AUCs for the best models (e.g., LightGBM and Ensembles) ranging narrowly from 0.67 to 0.77. ROC AUCs trended slightly lower in the lowest quantile (SESq1) and were the highest in SESq 5; moreover, ROC AUCs were almost identical in SESq 2-4. The Framingham’s AUCs were generally lower across SESqs (range 0.66–0.70), and showed no systematic trend. ML models showed slight improvements in PR AUC and F1 score, particularly in mid-to-high SESqs, though prevalence of hypertension was also higher (up to 14.9% in SESq 2). No model showed gross underperformance in any SES quartile, and differences between groups did not exceed 0.1 ROC AUC in the best models (LightGBM and ensembles). For a complete overview, see Table 6 of the S1 file. Additionally, all models’ ROC AUC (Figure 17), PR AUC (Figure 20), Brier (Figure 23), F1 (Figure 26), precision (Figure 29), and recall (Figure 32) plots by SESq can be found in the S2 file.

When we stratified entrants by educational level, models maintained moderate-to-high discrimination across most categories, with the highest AUCs in the largest and least educated group (level 1: n=1166, up to 0.729 for meta ensemble) and the most educated group (level 5, n=85, up to 0.808 for voting ensemble). However, confidence intervals for the smallest subgroups (level 4) were wide, due to the limited sample size (n=33). The ML models in the most educated participants’ group generally had the highest discrimination power (ROC AUC 0.80 in voting and weighted ensembles). The full Table of results is available in Table 7 of the S1 file. In addition, all models’ ROC AUCs (Figure 18), PR AUC (Figure 21), Brier (Figure 24), F1 (Figure 27), precision (Figure 30), and recall (Figure 33) plots by education can be found in the S2 file.

In all subgroups examined, ML models, particularly ensembles, offered improved discrimination compared to the Framingham score.

### Interpretability and Variable Importance

To avoid black-box concerns and have explainable ML models, SHAP and permutation importance analyses were performed on base models simultaneously (Figure 3). Our SHAP analysis showed that SBP was the most critical predictor in all base models; furthermore, in many models, the mean SHAP value of SBP exceeded 0.4. The second and third important predictors were age and BMI. Surprisingly, wrist circumference was among the most important features and in some models even the fourth predictor. Beeswarm plots of the base models can be found in Figure 34 of the S2 file. The permutation importance analysis indicated similar results, and SBP, age, and BMI were among the top predictor features (Figure 3b).

### Learning curves

To assess whether increasing training data improves model performance or not, we generated learning curves for each base model using the internal cross-validation data (Figure 35 of the S2 file). For each model, the ROC AUC was measured across incremental training set sizes. The train curves and the cross-validation curves for all models converged, with both training and cross-validation ROC AUC scores showing minimal change as the training set size increased.

The minimal change suggests that model performance has reached a plateau and is unlikely to benefit from additional data in the internal cohort. Consequently, the observed performance is likely limited by either model capacity or the limited available features in the dataset, rather than insufficient sample size

### Case vignette, Counterfactual analysis, and Bootstrap PI

Clinical risk and ML models are often opaque at the patient level. To provide deep-dive individual-level insights, we assessed ML models against FRS at the individual level. We selected 10 representative internal-test-set patients (5 with incident HTN, five without), all with BMI ≥ 20 and SBP ≥ 90, which reflects typical real-world phenotypes. For each representative patient, initially, we calculated the risk of HTN incident according to all base models, ensembles, and the Framingham rule, then we quantified the uncertainty in risk prediction for each patient in each model via 200 times resampling of the training set (bootstrapped 95% prediction intervals). According to the permutation and SHAP analyses, three features, namely age, BMI, and SBP, are consistently among the most important predictive variables. BMI and SBP are modifiable; therefore, to address the what-if questions, we performed counterfactual analysis (see S4 CSV table). We were specifically determined to answer whether we modify (lower) BMI and SBP in each patient when their HTN incident risk drops below a 0.2 threshold according to each model. For each model, we iteratively decreased BMI (from original to -4 units) and SBP (to -20 mmHg) and then plotted the HTN risk trajectory. Lastly, we compared predicted risks from major base and ensemble models for each patient using spider plots. Figure 4 shows the counterfactual analysis and bootstrap PI of two selected models (voting ensemble and Framingham); furthermore, Figure 4 also demonstrates the radar plots showing the risk of two discussed cases (patients 696 and 7976) predicted by these six models. Comprehensive counterfactual plots showing BMI and SBP risk trajectories for all 10 representative patients across all models, along with corresponding bootstrap 95% PI plots, detailed tables of prediction intervals, and radar plots comparing risk predictions for all patients, are provided in the S3 file.

At first glance, we identified heterogeneity across models. For individual patients, the original predicted probability of incident hypertension can span a wide range, even when trained on the same data. For example, patient 560’s original risk ranges from ∼0.17 (SVM) up to ∼0.66 (GB). Furthermore, ensemble risks often fall between the most conservative and most aggressive base models, showcasing their smoothing nature. Additionally, Framingham never predicted the risk more than 0.16, indicating its incompetence compared with ML models. Moreover, we found out that the threshold for low risk is not easily reached for many. For most of the positive outcome patients (HTN incident), even substantial reductions in BMI (e.g., -4 units) or in SBP (e.g., -20 mmHg) often do not bring the predicted risk below 0.2 in many models. For example, Patient 696’s risk in most models did not fall below 0.5 even with aggressive risk factor lowering. Some models exhibited step changes, especially decision-tree-based models and Framingham, indicating their categorical or bin-based logic. We also recognized that SBP reductions are often more potent than BMI; in several patients, lowering SBP results in larger risk reductions than BMI (see patients 7976 & 5891).

By reviewing counterfactual plots, it is evident that many ML models capture nuances unlike Framingham; most ML models recognize continuous small changes in BMI or SBP, when risk is lowered incrementally, while Framingham and tree-based models only adjust at certain thresholds. Predictably, negative-outcome patients’ original risks were much lower; more models allowed risk to fall below 0.2 with relatively modest adjustments, sometimes without needing any BMI or SBP reduction; nonetheless, there are several patients whose predicted risk never drops below 0.2, no matter how much you lower BMI or SBP. This is important because such patients likely have other risk factors (age, family history, etc.) that are unmodifiable in the model, and may point to clinical intractability.

For illustrative purposes, we assume patient 696 is admitted, the original BMI is 32.70, and the SBP reading is 125 mmHg (S4 CSV table and Figure 4). According to voting-ensemble, our best-performing model, the risk of developing HTN is 0.67 (bootstrap 95% CI: 0.62-0.71), the predicted risk according to Framingham for this patient is 0.162 (bootstrap 95% CI: 0.159-0.169). Counterfactual analysis based on voting ensemble indicates that if BMI is lowered by four units, the risk is still above 0.60, but if we decrease SBP by 20 mmHg, the risk is ∼0.55.

Alternatively, according to Framingham, if BMI is lowered by four units, the risk becomes 0.127, and similarly, if SBP is reduced by 20 mmHg, the risk becomes 0.127, although the actual outcome of patient 696 is HTN incident. Another case (patient 7976) is also admitted; the BMI is 25.78, and the SBP reading indicates 105 mmHg; the actual outcome for this patient is negative HTN. Voting ensemble’s prediction for HTN incidence for this patient is 0.25 (bootstrap 95% CI: 0.18-0.32). Framingham’s risk prediction for this case is 0.043 (bootstrap 95% CI: 0.039-0.046). The counterfactual analysis based on voting ensemble indicates that if the patient lowers their BMI by four units, the risk falls below the 0.2 threshold (0.17), and if SBP is reduced by 20 mmHg, the risk also falls below the 0.2 threshold (0.14). Nonetheless, the counterfactual analysis based on Framingham indicates that if BMI is lowered by four units, the risk becomes 0.033, and if SBP is reduced by 20 mmHg, the risk stays the same at 0.043. The complete table and all plots for the case vignette, counterfactual, radar plots, and bootstrap PI can be seen in the S3 file.

## Discussion

In this study, we conducted the most comprehensive ML approach for predicting HTN incidence. We evaluated all major ML families in predicting HTN incidence and benchmarked them against a clinical risk score (FRS). Our analysis showed that all ensembles and LightGBM outperform FRS ROC AUC-wise significantly (95% CIs did not cross 0). We externally validated our models and even created reduced models with similar feature mapping to external models for head-to-head comparison between internal and external models. Furthermore, we reported all major metrics in all models (ROC AUC, PR AUC, precision, recall, and Brier score) with their respective 95% CIs. We provided calibration curves across all internal and external models, along with their DCA. Although most models were overestimating the risk and DCAs mainly were flat, for the sake of transparency, we provided all the calibration and DCA curves.

Fairness of the ML models and their performance in different subgroups is often ignored; nonetheless, we conducted a comprehensive fairness gap analysis in males and females and showed that our models are fair to males and females. Moreover, our subgroup analysis in participants with different educational backgrounds, different socioeconomic statuses (SES), and in males and females indicated that, almost always, there are no significant drops in ROC AUC across various subgroups. For maximum interpretability and avoidance of black-box concerns, we simultaneously conducted SHAP and permutation importance analyses, both indicating the same important predictive features. Our SHAP and permutation analyses showed that wrist circumference is an important feature for HTN prediction. A recent study by Stankute et. al. also concluded that wrist circumference may be a new predictor of HTN consistent with our findings (11). To assess whether our sample size (∼8000 internal cohort) was enough or not, we drew learning curves of base models and classic voting and stacking ensembles. All curves suggested that after ∼2300 samples, ROC AUC becomes stable, thus indicating our sample size passes the minimum required size by a far margin. Lastly, while we were inspecting the current ML in medicine literature, we recognized that individualized ML is highly neglected. As a result, we designed a case vignette analysis comprising 10 real random test-set patients and calculated their HTN incident risk and its 95% CI using bootstrap PI, then we wanted to answer common what-if questions in those patients. After all, all the insights should be translated and applied in clinical practice; therefore, we chose two major HTN risk factors (BMI & SBP) and asked what if we lower BMI by four units and SBP by 20 mmHg? How would the risk change in these 10 patients? We calculated all necessary analyses and drew the plots (see S3 file). The comprehensive counterfactual analysis table can be accessed in the supplementary S4 CSV file. We concluded that although BMI & SBP are among the most important HTN predictors, lowering them does not necessarily reduce the HTN risk substantially, indicating there are numerous risk factors in HTN incidence, and none have a tremendous effect and predictive power.

The field of ML approaches in HTN is extremely underrepresented, and there are several challenges the field is facing, namely methodological flaws, lack of reproducibility, and lack of heterogeneity in the datasets and consequently lack of generalizability; furthermore, almost all prior studies fail to address these challenges properly (12). Confidently, we move the field forward by addressing all common issues in the field. For example, our pipeline presents one of the most comprehensive and modern ML in medicine pipelines; the code is fully reproducible and modular, which, with minimal modification, could be used for other ML prediction studies. Our used datasets (RaNCD and NHANES) are entirely separated; the health system, timeline, and even continents are different, ensuring models’ generalizability. A study conducted by Cheng et. al. on nearly 2 million Chinese participants utilized ML models to predict HTN incidence; their best model, CatBoost, had an ROC AUC of 0.888 (95% CI 0.886-0.889). Using SHAP analysis, they found that age, SBP, and BMI are the top HTN predictors (13). Nonetheless, their study lacked external validation making their models, performance prone to overfitting; furthermore, essential aspects that assure ML reliability including individualized ML (case vignettes), clinical benchmark (e.g., Framingham), learning curves, subgroup and fairness analyses were absent in their study, areas were tried to advance the field by ensuring that our models are fair, transparent, explainable, and clinically benchmarked.

The best-performing main internal models (voting & weighted ensembles) outperformed the best-performing reduced models (reduced voting & weighted ensembles) by the smallest margin (∼0.006 ROC AUC). The tiny difference adds credibility to the performance of external validation models and their results. Unlike the typical expectation of performance reduction on external validation datasets, our models showed robust generalizability, with the top-performing external models outperforming their corresponding main internal models. For instance, the voting and weighted ensemble models achieved substantially higher ROC AUCs on the external dataset (0.80 each) compared to their best internal performance (ROC AUC 0.72); furthermore, LR and SVM were the best performing on the external dataset (ROC AUC 0.82), reflecting an improvement of approximately 0.11. This trend was consistent for other key metrics, including PR AUC, F1 score, and Brier score. The mentioned results show our models’ predictive performance is not simply a result of overfitting to the internal data but rather reflects the strong underlying patterns generalizable across datasets. Several factors contribute to the mentioned outperformance; first, the label mix in NHANES differed from the internal dataset (approximately 1/3 compared to the internal cohort’s 1/10), which made NHANES a lot easier for the models compared to the internal dataset. Secondly, NHANES is one of the most respected datasets; there may be less noise in the NHANES dataset, and the data quality is high in such datasets.

The complete integration of clinical gold-standard comparison (FRS), a fully open and reproducible pipeline, and rigorous methodology are among our critical contributions. The online and public web calculator ensures that our models can be deployed in clinical practice from day zero. The narrative is suitable for both ML and clinical audiences. While absolute gains over FRS are moderate, ML models shone at the individual level; the counterfactual showed our ML models are sensitive to minimal risk factor modifications despite the cut-off approach in clinical risk scores. Future research should focus on multi-site validation, incorporate additional data sources, and novel ML algorithms.

The limitations include modest performance gains (ROC AUCs improved but not miraculously), limited Feature mapping between internal and external cohort (we were able to map only seven variables cross datasets), prevalence in the external dataset was compared to incidence in the internal dataset due to NHANES not providing incident cases, DCA and net benefit were minimal due to low baseline positives (11% internal), and class imbalance issues; we mitigated this issue via weighting/SMOTE, but it remains a source of uncertainty for some models and metrics.

In conclusion, our study shows that modern ML models, ensembles, and LightGBM substantially improve discrimination over FRS for incident hypertension prediction, with robust performance confirmed in both internal and external cohorts. The ML models consistently generalized across key demographic and socioeconomic subgroups, with comprehensive fairness and subgroup analyses showing minimal performance disparities. Our ML models are interpretable and clinically actionable via SHAP, permutation importance, and individualized counterfactual assessments. Furthermore, the models captured nuanced, patient-specific risk gradients unattainable by the categorical Framingham risk score. Our methodologically rigorous, transparent approach, fully open and reproducible code, and external validation address key barriers in the field and offer a robust foundation for translation into clinical practice.

### Ethical approval

Ethical approval for the original Ravansar Non-Communicable Disease (RaNCD) cohort study was granted by the Ethics Committee of Kermanshah University of Medical Sciences (approval code: KUMS.RE.1401.134). All procedures involving study participants were conducted in accordance with relevant institutional and national guidelines and regulations. Written and oral informed consent was obtained from all participants before enrollment in the cohort. This secondary analysis utilized anonymized data from the RaNCD study, with all identifying information removed before analysis. The NHANES dataset was publicly available.

## Code availability

The code used for analysis will be made available upon publication or acceptance.

## Authors’ contributions

Parsa Amirian:

Study design, data collection, data analysis, data interpretation, and writing the original manuscript.

Mahsa Zarpoosh:

Data collection, data interpretation, writing the original manuscript, revision, and figures

## Data Availability

The code and mock internal dataset and NHANES dataset used for the current analysis will be made available upon publication or acceptance.

